# The Current COVID-19 Spread Pattern in India

**DOI:** 10.1101/2020.06.03.20121210

**Authors:** Hemanta K. Baruah

## Abstract

In this article, we are going to show how to find out short term forecasts of the total number of COVID-19 cases in India in an easy way. Initially the spread of the disease was observably slow in India. Since the first week of May a highly nonlinear pattern has started to take shape. It can be observed that currently in India the spread pattern is nearly exponential. It can be seen further that the number of cases is still continuing to grow very fast. Therefore, instead of going for rigorous time series analysis, we may opt for looking at the data from a recent date downwards, and short term forecasts based on simple numerical analytical methods can be made accordingly.

## INTRODUCTION

To study the spread pattern of a pandemic such as COVID-19 there are standard mathematical methods based on solution of simultaneous differential equations as well as statistical techniques such as time series analysis. We have observed that after the time series graph starts to take a definite shape, we may try to use numerical analysis instead, one of the most classical branches of mathematics. In this kind of data made available at equal interval of time, it is obvious that there will be probabilistic error variables involved, and therefore a stochastic analysis is most appropriate in such a situation. However, randomness apart, numerical analytical methods are far simpler to deal with data that are being made available on a daily basis. To get an approximate mathematical expression to describe the data, one may take some recent data and try to find a polynomial expression using the forward difference operator. Indeed if we see that the graph concerned has started to take a very highly nonlinear shape, then we may simply try to fit an exponential pattern which would however be an approximation only, because the pattern cannot actually be exponential anyway, for if that happens that would mean that the pandemic would continue until everything is over. The pandemic has slowed down in various countries, and therefore a polynomial or an exponential curve cannot be the correct pattern, it can only be an approximate pattern valid for a short period.

In this article, we are going to show a very simple way to find short term forecasts of the COVID-19 spread in India. Not many works are available in the literature dealing with COVID-19 spread projections for India. Kumar *et. al.* [1] have discussed about forecasting of the spread using the auto-regressive integrated moving average (ARIMA) method of time series analysis. Poonia and Azad [2] have made forecasts for 22 April to 1 May, 2020, using the ARIMA method. It may be noted that the forecasts that they have put forward for those 10 days were very close to the realities. Azad and Poonia [3] thereafter extended their work to cover till 29 May.

Ghosh *et. al.* [4] ha*ve* used the Susceptible-Infectious-Susceptible (SIS) pandemic model to make certain predictions. The Center for Disease Dynamics, Economics & Policy (CDDEP), Washington, has produced a research report that includes predictions regarding infections from COVID-19 in India (Tseng *et. al.* [5]). This work was done during the very initial stage of the outbreak, before March 24.

Tomar and Gupta [6] have dealt with forecasting for India using data driven modeling. Tiwari *et. al.* [7] predicted that the pandemic was expected to peak in India between third and fourth week of April, and they predicted further that it would be controlled around the end of May. Their predictions have not actually been found to be correct.

Before going to discuss our methodology of forecasting, we would like to mention a few things about when the total number of cases started to grow in India. As per data published by Worldometers.info [8], the first COVID-19 cases were reported in India on 15 February, 2020; there were 3 cases reported. It may be noted that in the United States also the first cases were reported on the same date, 15 February, 2020; there were 15 cases reported. However the spread patterns were observably different in these two countries almost right from the beginning. On March 25 when the countrywide lockdown was imposed in India, there were just 657 cases. By April 15 when the second phase of lockdown was imposed, the number rose to 12370. By May 4 when the third phase of lockdown was imposed the number became 46437. We have observed that in India, approximately from the first week of May, the spread pattern started to show a highly nonlinear increasing trend.

## METHODOLOGY

In all the articles cited above, time series analysis and models for infectious diseases were used to make forecasts. Our standpoint is that we can make the process of forecasting quite simple if we can find that an approximate mathematical pattern is being followed by the data from some specific point of time, because if that can be observed then we need not apply time series models or any other models for infectious diseases to find what should the total number of cases be in the next few days. However our approach can be used for short term forecasting only.

We are going to show here that a rough short term forecast can easily be made if we can assume a nearly exponential pattern of the spread. Let us suppose that *N* is the value of total number of cases on any given day *t.* Let us assume that *N* = exp *(a + bt), a,b* >0, *t* ≥0, where a and *b* are constants. Then for *x = log_e_N = a + bt,* the first order differences Δ*x* of *x* would be constant for equidistant *t.*

Now consider the time series data of total number of cases in India from a recent day downwards. We have taken the data for our analysis from Worldometers.info [8] published on June 2, 2020. We have observed that the data were nonlinear almost from the beginning, and are currently very highly nonlinear. If the data show that the values of Δ*x* are very nearly constant, then we can easily make short term forecasts based on the recent observations assuming nearly exponential pattern of growth. However, if Δ*x* is not even nearly constant then this assumption of nearly exponential pattern of growth cannot be made, and in that kind of a situation we would go for a polynomial fit using forward difference interpolation. In what follows, we are going to check whether we can assume a nearly exponential pattern of growth in India at present.

In Table-1, we are showing values of *N* and Δ*x* for India from May 24 to May 11 downwards. It can be seen that from May 24 downwards to May 11, Δ*x* can be said to be nearly constant, and hence we can make an assumption of near exponential pattern of growth of *N* from May 11 to May 24 at least. We have checked that the values of were following a decreasing trend earlier to our start date of May 11. This is the reason why we cannot go for long term forecasts using our method. This is also the reason why our forecasts would be slight overestimations. It can be seen that the values of Δ*x* during these 14 days were around 0.05. Indeed the average of the values of Δ*x* comes out to be 0.051716.

**Table-1:** Total number of cases in India from May 11 to May 24.

| Date | $N$ | $X$ | $\Delta x$ |
| --- | --- | --- | --- |
| May 24 | 138536 | 11.838885 | 0.05270 |
| May 23 | 131423 | 11.786176 | 0.05175 |
| May 22 | 124794 | 11.734419 | 0.05406 |
| May 21 | 118226 | 11.680353 | 0.05384 |
| May 20 | 112028 | 11.626504 | 0.05083 |
| May 19 | 106475 | 11.575665 | 0.05946 |
| May 18 | 100328 | 11.516200 | 0.04724 |
| May 17 | 95698 | 11.468952 | 0.05421 |
| May 16 | 90648 | 11.414739 | 0.05515 |
| May 15 | 85784 | 11.359587 | 0.04515 |
| May 14 | 81997 | 11.314437 | 0.04926 |
| May 13 | 78055 | 11.265168 | 0.04941 |
| May 12 | 74292 | 11.215758 | 0.04859 |
| May 11 | 70768 | 11.167162 | 0.05231 |

From Table-2, it can be seen that from May 25 onwards the values of have started to show a slight decrease in comparison to the earlier 14 days. The average has come down to 0.045584 for the 7 days from May 25 to May 31. As the values of Δ*x* are still reducing, we can say that if we forecast the values of the total number of cases taking May 31 as base and Δ*x* = 0.045, we can go forward to make short term forecasts which will however be slight overestimations of the realities.

**Table-2:** Total number of cases in India from May 25 to May 31.

| Date | $N$ | $X$ | $\Delta x$ |
| --- | --- | --- | --- |
| May 31 | 190609 | 12.157979 | 0.04716 |
| May 30 | 181827 | 12.110810 | 0.04692 |
| May 29 | 173491 | 12.063881 | 0.04784 |
| May 28 | 165386 | 12.016037 | 0.04514 |
| May 27 | 158086 | 11.970894 | 0.04723 |
| May 26 | 150793 | 11.923663 | 0.03951 |
| May 25 | 144950 | 11.884144 | 0.04525 |

For that we shall follow the following procedure. On June 1, the value of *N* should approximately be equal to exp (12.157979 + 0.045) = 199382 at most. Here 12.157979 is the value of *x* for May 31. In the same way, the value of *N* for June 2 should be exp (12.157979 + 2 x 0.045) = 208559 at most. In Table-3 we have shown forecasts from June 1 to June 7.

**Table-3:** Forecasts for India from June 1 to June 7.

| Dates | Forecasts |
| --- | --- |
| June 1 | 199382 |
| June 2 | 208559 |
| June 3 | 218159 |
| June 4 | 228200 |
| June 5 | 238704 |
| June 6 | 249691 |
| June 7 | 261183 |

After a few days, we may check the average value of for another short period prior to that and the short term forecasting may be continued. Our proposed method is neither highly mathematical nor statistical, but it does return workable forecasts in a very simple way. We have used this method to find an approximate pattern of COVID-19 spread outside China, and we have seen that the method gives slightly overestimated forecasts [9] because in certain countries the spread has already started to show a pattern.

## CONCLUSIONS

For immediate planning, the policy makers in India may be interested to forecast the total number of COVID-19 cases. The spread pattern in India is very nearly exponential already. Very short term forecasts can therefore be made in a simple way because we have found that the COVID-19 spread pattern in the country is nearly exponential. To check whether it is true, because the pattern might suddenly change, we need to look at the time series data from a recent date downwards, and rough but workable short term forecasts can be found easily.

## Data Availability

The data were taken from Worldometers.info.

https://www.worldometers.info/coronavirus/coronavirus-cases/

## Notes

### Competing Interest Statement

The authors have declared no competing interest.

### Clinical Trial

This work is not based on clinical trials.

### Funding Statement

This work is not supported by any funding agency.

### Author Declarations

This work does not need any approval of the IRB/oversight body.

## REFERENCES

1. P. Kumar, R. K. Singh, C. Nanda. et. al. Forecasting COVID-19 impact in India using pandemic waves nonlinear growth models, medRxiv preprint doi: https://doi.org/10.1101/2020.03.30.2004703.

2. N. Poonia, S. Azad, Short term forecasts of COVID-19 spread across Indian States until May 1, 2020, arXiv: 2004.13538v2[q.bio.PE ].

3. S. Azad, N. Poonia, Short term forecasts of COVID-19 spread across Indian States until 29 May, 2020, under the worst case scenario, Preprints 202000491. https://doi.org/10.20944/preprints202004.0491.v1

4. P. Ghosh, R. Ghosh, B. Chakraborty, COVID-19 in India: State-wise analysis and prediction, medRxiv preprint, doi: https://doi.org/10.1101/2020.04.24.20077792.

5. K. Tseng, E. Schueller, G. Kapoor, et. al. Center for Disease Dynamics, Economics & Policy, Washington, DC, COVID-19 for India updates, March 24, 2020.

6. A. Tomar, N. Gupta, Prediction for the spread of COVID-19 in India and effectiveness of preventive measures, Science of the Total Environment, 728 (2020) 138762. https://doi.org/10.1016/j.scitotenv.2020.13762.

7. S. Tiwari, S. Kumar, K. Guleria, Outbreak trends of coronavirus disease-2019 in India: A prediction, Disaster Med Public Health Prep. (2020) 1–6. doi: 10.1017/dmp.2020.115.

8. Worldometers.info. Total coronavirus cases in India, Publishing Date: June 2, 2020. Place of Publication: Dover, Delaware, U. S. A.

9. H. K. Baruah, medRxiv preprint doi: https://doi.org/10.1101/2020.05.24.20112292. Posted on May 30, 2020.

